# Incidence and predictors of relapse among children with nephrotic syndrome at Assosa zone hospitals, Benishangul Gumuz Region, Northwest Ethiopia, 2022

**DOI:** 10.1101/2022.10.20.22281301

**Authors:** Yegoraw Gashaw, Berhan Tekeba, Bewuketu Terefe, Nega Tezera, Seid Wodajo

## Abstract

**Background:** Relapse is the major problem in children with nephrotic syndrome and leads to a substantial burden on the patient and family worldwide, particularly in resource-limited countries like Ethiopia. However, little is known about the incidence of relapse and its predictors among children with nephrotic syndrome in the study area.

**Methods:** An institution-based retrospective follow-up study was conducted among 354 randomly selected nephrotic syndrome patients admitted from April 2017 to March 2022. Data entry was carried out using Epi-data manager version 4.6.0.6 and Stata software version 14 for data cleaning and analysis. Cox-proportional hazard models were used to identify predictors of relapse. Any variable with a p-value < 0.25 in the bivariable was taken into the multivariable analysis, and then the association and statistical significance were declared at P≤ 0.05.

**Results:** More than half, 55.8% (82/147), of relapses were recorded in the first six months of follow-up. The incidence of relapse was 82.3 per 1000 child-month-observations, with an overall risk of 1785.9 child-month-observations. In children with nephrotic syndrome, the presence of wasting malnutrition [AHR = 1.93, 95% CI 1.28–2.90], acute respiratory tract infections [AHR = 1.79, 95% CI 1.19–2.71], elevated triglyceride levels [AHR = 2.74, 95% CI 1.48–5.07], and low serum albumin levels [AHR = 4.34, 95% CI 22.18–8.64] were predictors of relapse.

**Conclusion and Recommendations:** The incidence of relapse among nephrotic syndrome patients was high. The independent predictors of relapse in children with nephrotic syndrome were the presence of acute respiratory tract infections, wasting malnutrition at admission, low serum albumin levels, and elevated serum triglyceride levels. Therefore, intervention to reduce and control earlier relapse should focus on preventing relapse-related complications.

## Introduction

Nephrotic syndrome is a chronic glomerular disease that presents in children as relapsing-remitting courses, with an incidence of 1.15 to 16.9 per 1000 children (1). Relapse in children with nephrotic syndrome is the reappearance of proteinuria for at least three consecutive days following achievement of remission (1, 2). Although nephrotic syndrome can affect children of any age, from infancy through adolescence (3), the risk of relapse is highest in school-aged groups; nevertheless, the risk of relapse reduces in adolescents (4).

Relapse in nephrotic syndrome has been reported to be 71.9 % worldwide (5). 80-90% of children experience one or more relapses after the onset of nephrotic syndrome and about 50% will develop frequent relapses (6). However, there is substantial variability in the occurrence of relapse across the countries. A shred of recent evidence in France (79%), Australia (80%), and Bangladesh (70.1%) indicates that there could be a higher relapse rate among children with nephrotic syndrome (6-8). In Abuja, Nigeria 63.3% of children experienced a relapse (9).

Despite several attempts to reduce its recurrence, relapse continues to be the leading cause of complications such as infections, thrombosis, dyslipidemia, malnutrition, and lower quality of life (10). In low income-countries, relapse-related complications remain high, which could be disproportionately affected because of less advanced technology for early diagnosis, prevention, and management of nephrotic relapses (11). The increasing rate of non-communicable disease in Africa makes understanding possible risk factors of relapse among children with nephrotic syndrome. However, relapse of nephrotic syndrome can lead to increase the risk of developing chronic kidney disease that confers significant morbidity and mortality (12).

In Ethiopia, there are limited studies on the incidence of relapse and its predictive factors among children with nephrotic syndrome. However, it was the second most common cause of pediatrics admission due to renal disease with the relapse rate range from 35.6% to 78% (13-15). As a result of the disease process, relapses have a tremendous impact on various biological activities associated with protein loss during the active phase, which may lead to increased comorbidities, systemic complications, financial burden, and lower quality of life (1, 16). Relapses that occur within the first year are an independent predictor of later subsequent relapse, and those that occur within the first six months of diagnosis are highly indicative of how the disease process will progress (17, 18). The common triggers for relapse in children with nephrotic syndrome includes infection, allergies(19), stress (20), and cessation of steroids (21).

Globally, various initiatives have been attempted to lower the risk of relapse in children with nephrotic syndromes, such as zinc supplementation, use of immunosuppressive agents, and daily administration of prednisolone therapy during episodes of upper respiratory tract infections (22-24)]. Apart from these initiatives, the issue continues to get attention from the general population. However, as far as the investigator searched, there is limited information on the incidence of relapse and its predictor factors among children with nephrotic syndrome in Ethiopia, particularly in the study area. Hence, this study aimed to assess the incidence of relapse and its predictors among children with nephrotic syndrome at Assosa zone hospitals, Benishangul Gumuz, Northwest Ethiopia.

## Materials and Methods

### Study design, period, and setting

An institution-based retrospective follow-up study was conducted among children with nephrotic syndrome from April 2017 to March 2022 at Assosa zone hospitals, Benishangul Gumuz, Northwest Ethiopia. Assosa zone is found in Benishangul Gumuz Regional State, 661 km far west from Addis Ababa, which is the capital city of Ethiopia, with approximately 439,786-projected population. It shares borders with Sudan in the northwest, the regional state of Amhara in the east, and West Oromia in the south. The zone have two public hospitals namely; Assosa general hospital and Menge primary hospital. Assosa general hospital serves people who come from Assosa zone, west Oromia, and South Sudan refugees, whereas Menge primary hospital serves people who come from Menge woreda and the nearby border of Sudan. They gives both outpatient and in-patient services like medical, surgical, pediatrics, ICUs, obstetric and gynecologic, emergency care in the zone.

### Populations

The source population of this study included all medical records of children below 15 years of age who had a follow-up with nephrotic syndrome at Assosa zone hospitals, and the medical records of children below 15 years of age who were diagnosed with nephrotic syndrome from April 2017 to March 2022 were the study population.

### Sample size determination and sampling procedures

For the first objective, we computed a single population proportion formula by considering the following assumptions: 95% CI, 5% margin of error, and a 35.6% proportion of relapse rate among patients with nephrotic syndrome. However, with a 10% incomplete rate, the final sample size was 389. For the second objective, we computed the sample size using the Cox proportional hazard model in Stata software Version 14 by considering the assumptions of the following predictors, such as undernutrition, low serum albumin levels, elevated triglyceride levels, and child’s residence. By considering the following assumptions Z = is the critical value of a standard normal distributed variable at 5% significance level = 1.96. Z = is the critical value of a standard normal distributed variable at 20% = 0.80. The probability of an event was 0.356 (14). Moreover, the probability of withdrawal is 0.05 and using an adjusted hazard ratio. Therefore, we used the largest sample size that was calculated for the first objective (N = 389). Then, a computer-generated simple random sampling technique was employed based on the child’s medical registration number to select the patient’s chart. The details of steps and procedures were described in the **figure 1** below.

**Figure 1:**
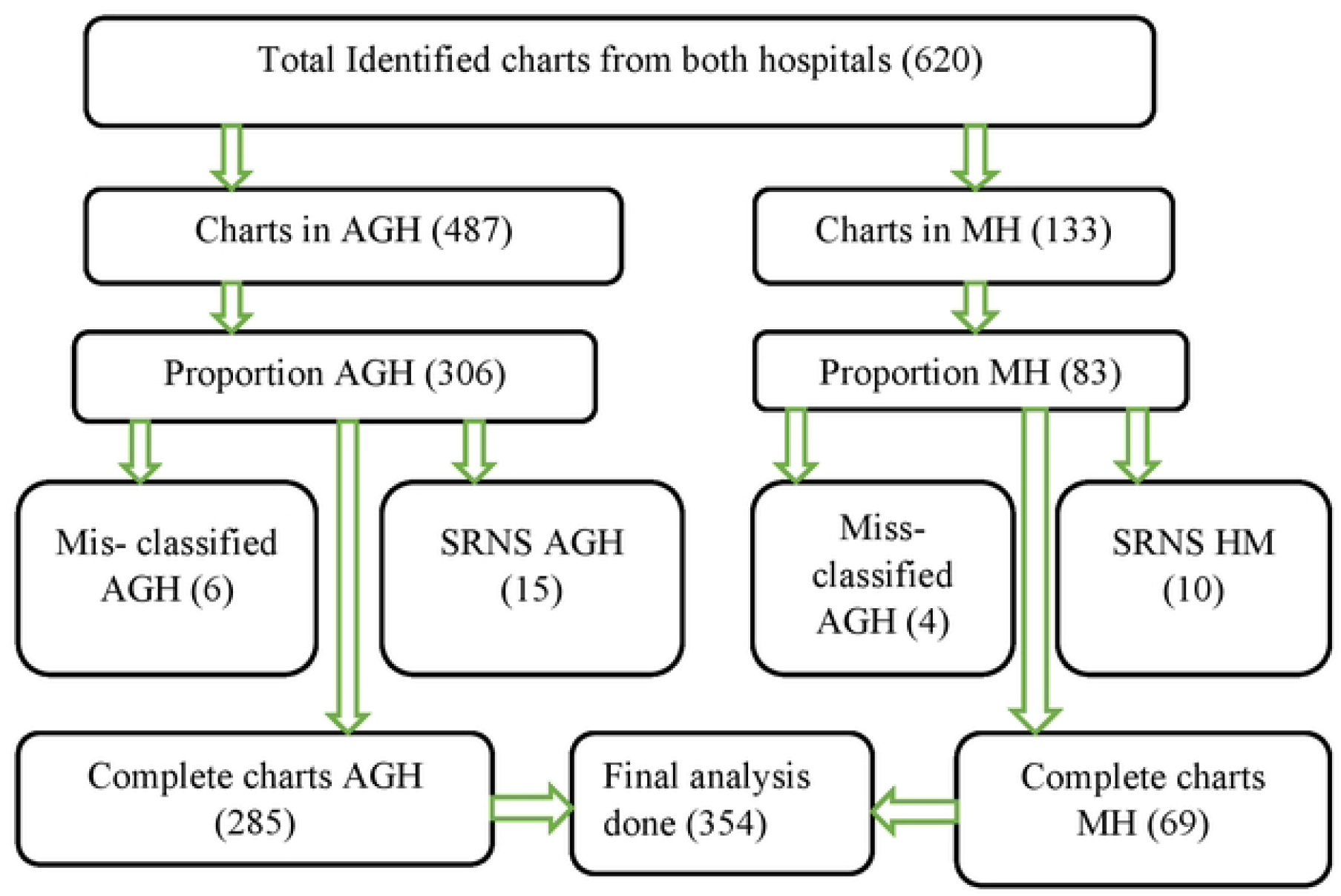
Schematic presentation of sampling procedure to assess the incidence and predictors of relapse among children with nephrotic syndrome in Assosa zone hospitals, Benishangul Gumuz, Northwest Ethiopia, 2022. NB: AHG: Assosa General Hospital, MPH: Menge Primary Hospital.

### Operational definitions and measurements

Events were children with nephrotic syndrome who developed their first relapse after they achieved remission from nephrotic syndrome. Children having less than 1+ proteinuria (proteinuria 40 mg/m^2^/hour) for three consecutive days in a week after completion of treatment were considered to have remission (1). In previously remitted patients, relapse was defined as the recurrence of proteinuria of ≥3+ for three consecutive days or 1+ for seven days in a week and declared when the patients’ records showed relapse and /or when the diagnostic criteria were fulfilled (1). Steroid sensitive nephrotic syndrome was defined as the achievement of complete remission within eight weeks of steroid therapy after initial presentation. Steroid-resistance was the absence of remission despite daily corticosteroid therapy (1). Censored were those cases that had never developed a relapse, transfer out, and lost to follow up during the follow-up. Survival time was measured from previous remission to the occurrence of the first relapse and the follow-up time of relapse was measured in months.

For children with nephrotic syndrome, malnutrition was classified as wasting in to severe acute malnutrition and moderate malnutrition whereas, and stunting according to the WHO z-score measurement scale. SAM is diagnosed if WHO z-score of weight-for- height (WFH) < - 3 or mid-upper arm circumference (MUAC) < 115mm, or there is bilateral pitting edema and/ physician diagnosis or, MAM if WFH between -3 and -2 z-score without pitting edema (25) and physician diagnosis. BMI for age (BAZ) below - 2 Z score will be considered to diagnose wasting for children above six years. Children were declared to have hematuria based on the diagnosis noted on their charts. The presence or absence of infection records on the patient’s charts plus their respected laboratory results.

### Data collection tools and procedures

The data available in the patient chart was checked first and the extraction checklist was prepared in English, which was adapted from the renal clinic follow-up registration book and by reviewing relevant - related literature. The tool is comprised of sociodemographic characteristics, baseline disease-related characteristics, baseline biochemical-related characteristics, and treatment-related characteristics. Charts were accessed based on their medical registration numbers. The baseline biochemical-related test results were taken from their last remission records. The beginning of chart review was from the date of last remission and the end time was the date of relapse, recovery, lost to follow-up, and transfer out. Four BSc degree nurses were recruited and the data was collected from April to May 2022. Throughout data collection, both the supervisor and the principal investigator did the supervision.

### Data processing and analysis

The Epi-data manager version 4.6.0.6 was used to collect data, which was then transferred to Stata software version 14 for cleaning, editing, coding, and analysis. The presence of outliers and missing values were checked before the analysis of the data. A Kaplan-Meier relapse curve was carried out to estimate survival failure probability, and log-rank tests were used to compare survival curves for the presence of a difference in the incidence of relapse among the groups. The statistical significance was declared by using an adjusted hazard ratio at 95% CI with a p-value less than 0.05 in the multivariable Cox proportional hazards regression model. Proportionality assumptions were tested using a global test with a value of P> χ2 =0.9053. The goodness of fit of the final model was checked by the Nelson–Alen cumulative hazard function against the Cox– Snell residual test.

### Data quality control

A pretest was conducted on twenty-three randomly selected medical charts at Kamashi primary hospital, which is not included in the present study. One-day training was given for both data collectors and supervisor on the extraction process and the way they were going to collect relevant information. The principal investigator and supervisor closely monitored the entire data collection process, checked the completeness and consistency of the collected data, and gave prompt feedback to the data collector. The collected data was crosschecked daily during and after data collection.

### Ethical consideration

Ethical approval was received from the school of nursing research ethical review committee on behalf of University of Gondar, institutional review board (IRB). Then, an official letter of cooperation was written to Assosa General Hospital and Menge Primary hospital from the school of nursing, department of pediatric and child health nursing on behalf of University of Gondar. Then, permission for data collection was obtained from each hospital’s administration on behalf of patients, renal clinic, and pediatric wards of each hospital. The information obtained was kept confidential and was only used for the study.

## Results

### Sociodemographic characteristics

The findings of this study revealed that 193 (54.5%) participants were male. Of those, 105 (54.5%) experienced relapse. About 202 (57.06%) children were diagnosed with nephrotic syndrome in the age range of four to eight years old, with a median age of seven years. The result also showed that about 55.1% of children came from rural areas; of those, 41% (80/195) of children with nephrotic syndrome developed relapse. However, none of the study participants had a known family history of renal disease (**Table 1**).

**Table 1:** Sociodemographic characteristics of children with nephrotic relapse admitted at Assosa zone hospitals, Benishangul Gumuz Region, Northwest Ethiopia; 2022 (N=354).

| Variables | Category | Frequency (n) | Status |  | Percent (%) |
| --- | --- | --- | --- | --- | --- |
|  |  |  | Event N (%) | Censored N (%) |  |
| Age at onset (years) | 1-4 | 72 | 25(34.72) | 47(65.28) | 20.34 |
|  | 4-8 | 202 | 102(50.5) | 100(49.5) | 57.06 |
|  | 8-15 | 80 | 20(25) | 60(75) | 22.60 |
| Sex | Male | 193 | 105(54.4) | 88(45.6) | 54.5 |
|  | Female | 161 | 42(26.1) | 119(73.9) | 45.5 |
| Residence | Rural | 195 | 80(41) | 115(59) | 55.1 |
|  | Urban | 159 | 67(42.1) | 92(57.9) | 44.9 |

### Baseline disease-related characteristics of study participants

More than half (53.11%) of study participants had an acute respiratory tract infection, of which 55.3% of children developed a relapse. Moreover, among 250 (70.62%) children with urinary tract infection cases, 45.2% experienced a relapse. Regarding nutritional status, almost 93% of children with wasting malnutrition developed relapse of nephrotic syndrome. Meanwhile, 122 (58.4%) children with hypertension experienced a relapse of nephrotic syndrome, and nearly 49% of children with a history of allergy had developed a relapse (**Table 2**).

**Table 2:** Disease-related characteristics of children with nephrotic syndrome in Assosa zone hospitals Benishangul Gumuz, Northwest Ethiopia, 2022 (N=354).

| Variables |  | Category | Frequency<br>(N) | Status |  | Percent (%) |
| --- | --- | --- | --- | --- | --- | --- |
|  |  |  |  | Event N (%) | Censored N (%) |  |
| Infection | UTIs | Yes | 250 | 113(45.2) | 137(54.8) | 70.62 |
|  |  | No | 104 | 34(32.7) | 70(67.3) | 29.38 |
|  | ARTIs | Yes | 188 | 104(55.3) | 84(44.7) | 53.11 |
|  |  | No | 166 | 43(25.9) | 123(74.1) | 46.89 |
| Nutritional<br>status | Wasting | Yes | 96 | 89(92.7) | 7(7.3) | 27.12 |
|  |  | No | 258 | 58(22.5) | 200(77.5) | 72.88 |
|  | Stunting | Yes | 32 | 21(65.63) | 11(34.37) | 9.04 |
|  |  | No | 322 | 126(39.1) | 196(60.9) | 90.96 |
| Hypertension |  | Yes | 209 | 122(58.4) | 87(41.6) | 59.04 |
|  |  | No | 145 | 25(17.2) | 120(82.8) | 40.96 |
| Allergic disease |  | Yes | 205 | 100(48.8) | 105(51.2) | 57.9 |
|  |  | No | 149 | 47(31.5) | 102(68.5) | 42.1 |
| Time to remission in<br>days |  | < 7 | 88 | 35(40) | 53(60) | 24.9 |
|  |  | 8- 14 | 172 | 67(39) | 105(61) | 48.5 |
|  |  | >14 | 94 | 45(48) | 49(52) | 26.5 |

### Treatment-related characteristics of study participants

Almost 98% of the study participants were given prednisolone medication, of which 140 (40.5%) children experienced a relapse of nephrotic syndrome. Nearly half (48.5%) of the study participants on steroid therapy achieved remission within two weeks of treatment completion. None of the study participants had a history of zinc supplementation for respiratory tract infections in children with nephrotic syndrome (**Table 3**).

**Table 3.** Treatment-related characteristics of children with nephrotic syndrome in Assosa zone hospitals Benishangul Gumuz, Northwest Ethiopia, 2022 (N=354).

| Variables | Category | Frequency<br>(N) | Status |  | Percent (%) |
| --- | --- | --- | --- | --- | --- |
|  |  |  | Event N (%) | Censored N (%) |  |
| Prednisolone<br>received | Yes | 346 | 140(40.5) | 206(59.5) | 97.74 |
|  | No | 8 | 7(87.5) | 1(12.5) | 2.26 |
| Duration of<br>treatment (week) | ≤4 | 181 | 92(51) | 89(49) | 51.1 |
|  | 4-8 | 173 | 55(32) | 118(68) | 48.9 |

### Biochemical-related characteristics of the study participants

One hundred fifteen study participants had a serum protein level of below 3g/dl at previous remission, of which 76.5% (88/115) developed relapse of nephrotic syndrome. Moreover, 257 (72.6%) children had a serum albumin level of less than or equal to 1.5 g/dl, of which 53.3% experienced relapse. Among children having high triglycerides, 52% developed relapse of nephrotic syndrome. Furthermore, hundred ninety-nine (56.2%) participants had hematuria in which 33.3% (22/66) of them were found to be relapsed (**Table 4**).

**Table 4:**
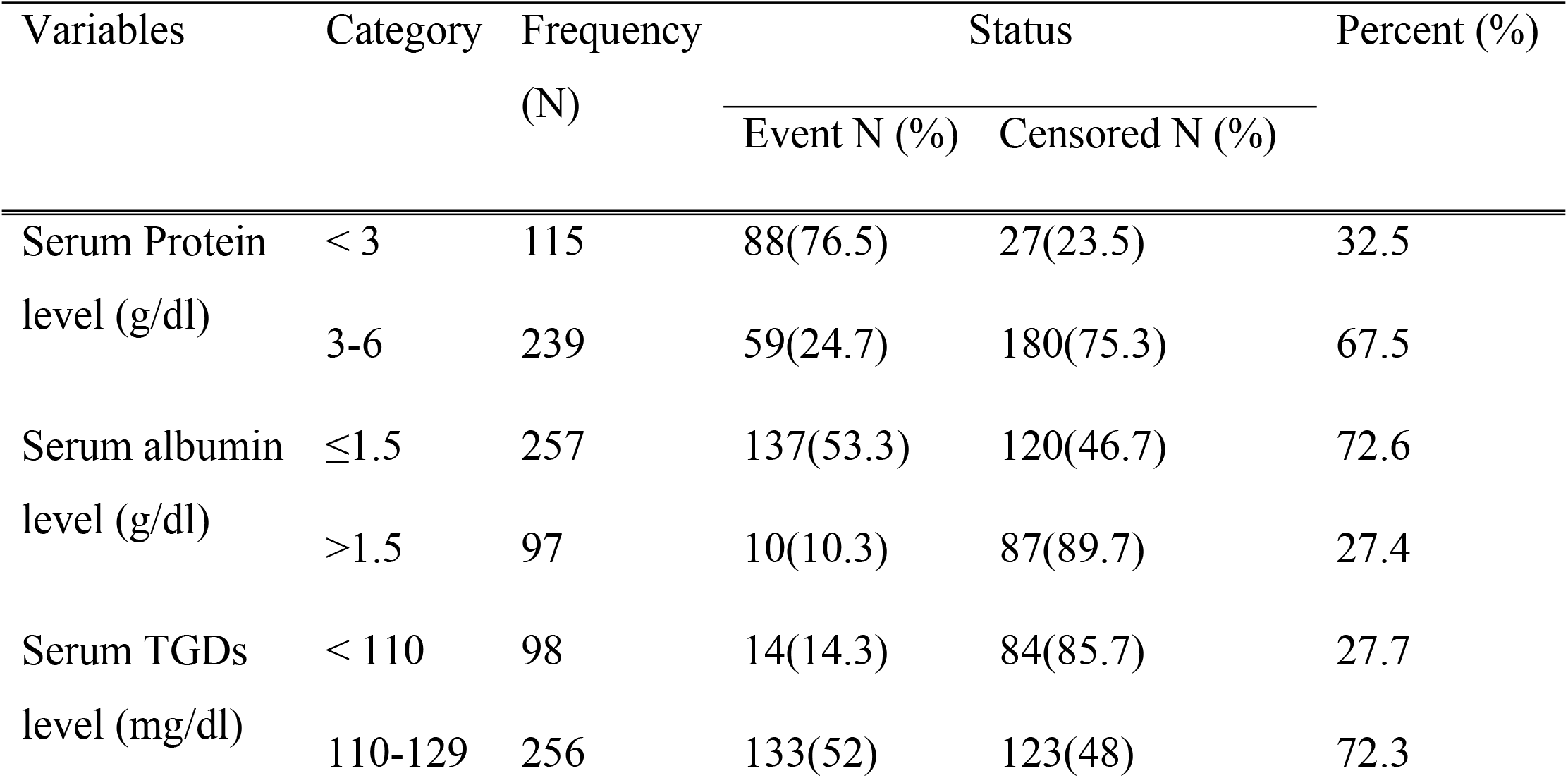
Biochemical-related characteristics of children with nephrotic syndrome in Assosa zone hospitals Benishangul Gumuz, Northwest Ethiopia, 2022 (N=354).

### Incidence of relapse

During the follow-up period, the incidence of relapse among children with nephrotic syndrome was 82.3 (95% CI: 70.03–96.75) per 1000 child-months of observation, with the total time at risk of 1785.9. The study participants were followed-up for a minimum of 1 month and a maximum of 12 months, with a median follow-up time of 6.2 months. The overall Kaplan-Meier failure function showed that the probability of relapse among children with nephrotic syndrome increased during the entire follow-up time. Children’s survival failure estimates varied depending on their serum albumin, triglyceride level, wasting malnutrition, and presence of acute respiratory tract infections at admission. From the months of five to nine, the curve tends to rise rapidly, implying that most children relapse within this period. The median follow-up time of the study participant was 6.2 months with median time to relapse of 5.2 months (**Figure 2**).

**Figure 2:**
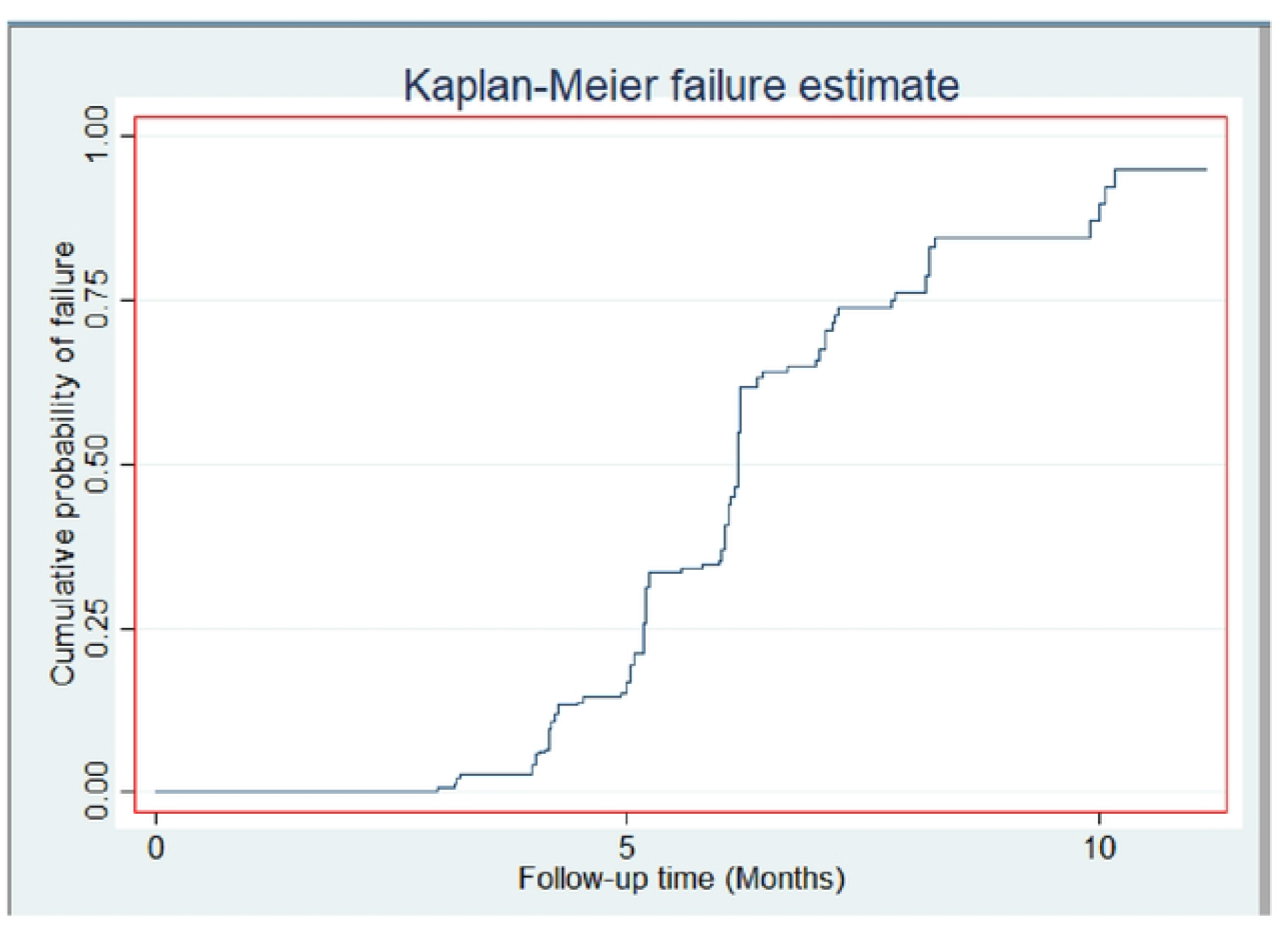
Overall Kaplan-Meier failure curve of children with nephrotic syndrome followed-up at Assosa zone hospitals, Benishangul Gumuz, Western Ethiopia, 2022 (n=354).

### Predictors of relapse

In the final Cox-proportional hazard analysis having wasting malnutrition, acute respiratory tract infection, high triglyceride levels, and low serum albumin levels have been found to be independent predictors of relapse in children with nephrotic syndrome. When all other variables were held constant, children who had an acute respiratory tract infection at admission had a 1.79 times higher risk of developing a nephrotic syndrome relapse [AHR = 1.79, 95% CI: 1.19-2.71] than those who did not have an upper respiratory tract infection. Furthermore, children with wasting malnutrition at admission had a 1.93-fold higher risk of relapse compared to children who were well nourished [AHR = 1.93, 95% CI (1.28-2.90)]. Likewise, children who had high triglyceride levels [>110mg/dl] increased the hazard of relapse by 2.74 times as compared with those who had low serum triglyceride levels [AHR: 2.74; 95% CI: 1.48–5.07]. Furthermore, children with low albumin levels [1.5g/dl] at baseline were 4.34 times more likely to develop relapse than those with serum albumin levels > 1.5g/dl [AHR = 4.34, (95% CI: 2.18-8.64) (**Table 5**).

**Table 5.** Bi-variable and multivariable cox regression analysis for predictors of relapse among children with nephrotic syndrome followed-up at Assosa zone hospitals from April 2017 to March 2022, Benishangul Gumuz, western Ethiopia, 2022. (N= 354).

| Variables | Category | Status |  | CHR | AHR |
| --- | --- | --- | --- | --- | --- |
|  |  | Event N (%) | Censored N (%) | (95% CI) | (95% CI) |
| Age at onset in years | 1-4 | 25(34.72) | 47 (65.28) | 0.46(0.15-1.40) | 1.49(0.745-2.98) |
|  | 4-8 | 102 (50.5) | 100 (49.5) | **2.82(1.025-7.73)\*** | 1.56(0.94-2.69) |
|  | 8-15 | 20(25) | 80 (75) | 1 | 1 |
| Sex | Male | 105 (54.4) | 88 (45.6) | **1.69(1.18-2.42)\*** | 1.146(0.78-1.68) |
|  | Female | 42 (26) | 119 (74) | 1 | 1 |
| Residence | Rural | 107 (54) | 91 (46) | **2.28(1.58-3.28)\*** | 1.33(0.89-1.98) |
|  | Urban | 24 (20) | 96 (80) | 1 | 1 |
| UTIs | Yes | 113 (45.2) | 137 (54.8) | **1.50(1.02-2.21)\*** | 1.25(0.79-1.99) |
|  | No | 55 (27) | 151 (73) | 1 | 1 |
| ARTIs | Yes | 55 (33) | 110 (67) | **2.33(1.63-3.32)\*** | **1.79(1.19-2.71)\*\*** |
|  | No | 92 (49) | 97 (51) | 1 | 1 |
| Wasted malnutrition | Yes | 56 (89) | 7 (11) | **2.40(1.74-3.33)\*** | **1.93(1.18-2.90)\*\*** |
|  | No | 91 (31) | 200 (69) | 1 | 1 |
| Stunted malnutrition | Yes | 15 (58) | 11 (42) | **1.95(1.22-3.10)\*** | 1.23(0.71-2.13) |
|  | No | 132 (40) | 196 (60) | 1 | 1 |
| Hypertension | Yes | 122 (58) | 87 (42) | **1.95(1.26-3.01)\*** | 1.05(0.65-1.69) |
|  | No | 25 (17) | 120 (83) | 1 | 1 |
| Allergic disease | Yes | 99 (49) | 105 (51) | **1.49(1.02-2.17)\*** | 1.26(0.86-1.85) |
|  | No | 25 (17) | 120 (68) | 1 | 1 |
| Duration of treatment (week). | ≤4 | 92(51) | 89(49) | **1.54(1.10-2.15)\*** | 1.03(0.7-1.5) |
|  | 4-8 | 55(31.8) | 118(68.2) | 1 |  |
| Serum protein level (g/dl) | ≤ 2 | 112 (89.6) | 13 (10.4) | **3.83(2.62-5.61)\*** | 1.38(0.94-2.03) |
|  | 3-6 | 35 (15) | 194 (85) | 1 | 1 |
| Serum albumin level (g/dl) | ≤1.5 | 137 (53) | 120 (47) | **5.75(3.02-10.95)\*** | **4.34(2.18-8.64)\*\*** |
|  | >1.5 | 10 (10) | 87 (90) | 1 | 1 |
| Serum TGDs level (mg/dl) | <110 | 142 (41) | 203 (59) | 1 | 1 |
|  | ≥110 | 5 (56) | 4 (44) | **1.22(0.50-2.98)\*** | **2.74(1.48-5.07)\*\*** |
| Hematuria | Yes | 23 (92) | 2 (8) | **1.57(1.00-2.45)\*** | 1.59(0.98-2.57) |
|  | No | 124 (38) | 205 (62) | 1 | 1 |

NB: \* significantly associated variables in bi-variable Cox-regression, \*\* significantly associated variables in multivariable Cox-regression, AHR: adjusted hazard ratio, CHR: crude hazard ratio

### Assessment of model adequacy

The overall Cox-residual Schoenfeld global test was checked for proportional hazard assumption and it was met (p-value = 0.9053). All covariates are met the proportional-hazard assumption. The goodness-of-fit was tested by Cox-Snell residuals. For the residual test, it was possible to conclude that the final model fits the data well, which means these residuals should have a standard censored exponential distribution with a hazard ratio. The hazard function follows the 45° line very closely (**Figure 3**).

**Figure 3.**
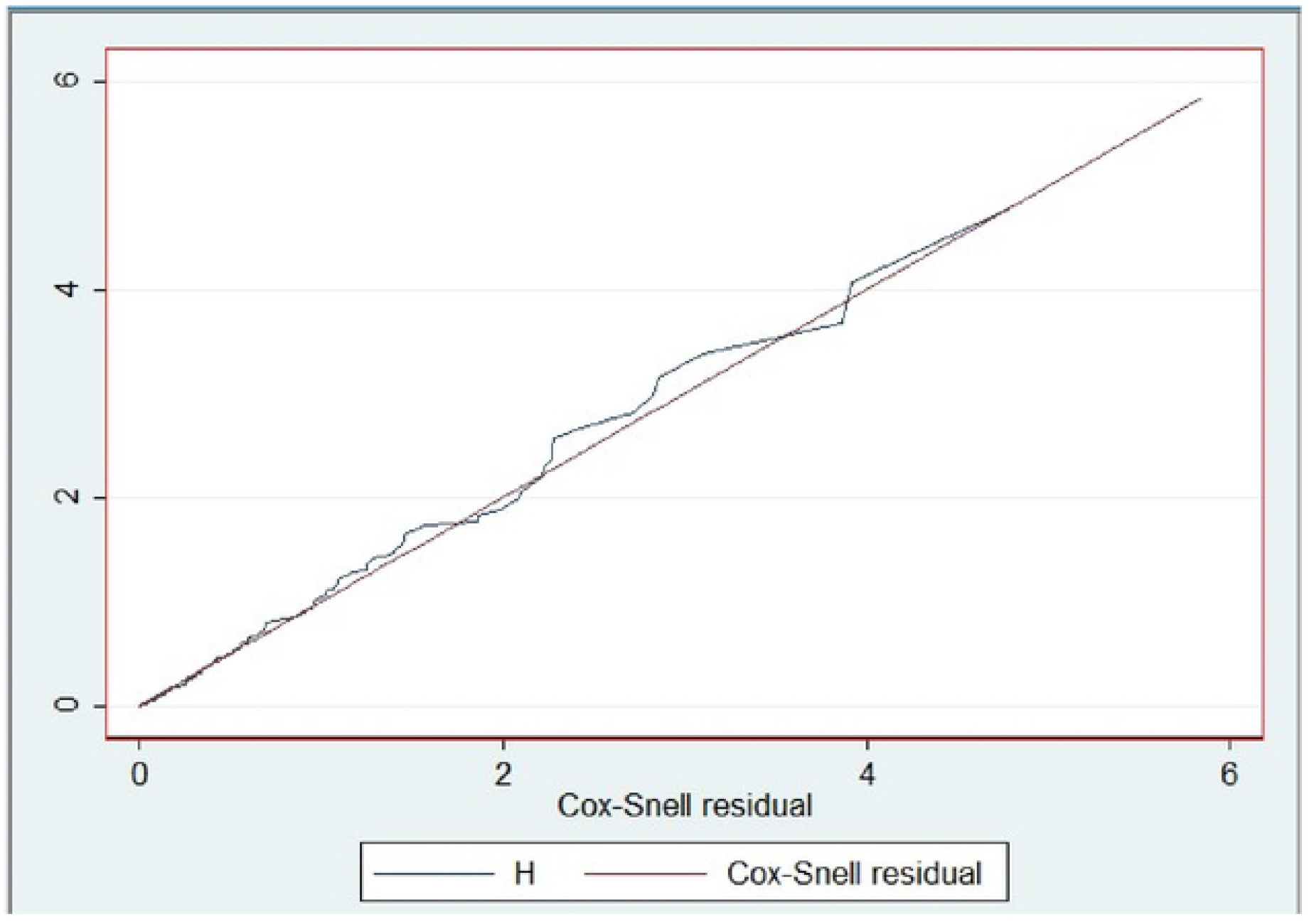
Cox-Snell residual test for children with nephrotic relapse followed-up at Assosa zone hospitals, Benishangul Gumuz, western Ethiopia, 2022 (N=354).

## Discussion

Relapse is one of the major health problem in children with nephrotic syndrome. The incidence rate of relapse was 82.3 per 1000 child month-observations. About 41.5% of children with nephrotic syndrome developed relapse. This finding was lower than the reported relapse rate of between 60% and 90% among initial responders of pediatric nephrotic patients (9). However, the relapse rate in this study was also lower than the 89.7%, 83%, and 79% relapse rates reported in Sudan, country of Gironde France, and population-based cohort in France (6, 26, 27). The possible explanation for this discrepancy could be due to difference among the study population, ethnicity, duration of follow-up, and age at onset of the disease. Moreover, it was lower compared with reports of a relapse rate ranging from 48.7% to 78% among children with steroid-sensitive nephrotic syndrome in Ethiopia (15, 28). This variation might be due to difference in sample size, study population, and presence of complication at diagnosis in the current study. However, the finding is higher than a study done in norther Ethiopia that was 35.6% (14). The possible discrepancy might be due to the difference in the age of the study population, limited availability of health services, and scarcity of essential laboratory parameter at peripheral health facilities in the current study. The finding of the current study was also lower than that of studies from Indonesia (56.3%) and India (2013) (59.3%). The possible reasons could be due to the difference in sociodemographic characteristics, sample size, and clinical factors of the study population in those studies.

This study found that the median time to relapse was 5.2 months. It was lower compared to previous studies done in Ethiopia, which was 16 months (14), France (7), and Nigeria (29) which were 11 and 8.3 months respectively. The possible discrepancy for this variation could be due to difference in follow-up times in those studies. However, it was higher than in studies done in India and Australia where the median time to relapse was 5 months (6). This might be due to a relatively higher incidence of infections, malnutrition, and limited access to healthcare services in the current study area.

The finding suggested that children who had acute respiratory tract infections at admission were about 1.79 times increased hazard of developing relapse than those who had no respiratory tract infections. This finding is in agreement with the study done in Indonesia (30) and Bangladesh (8). The similarity could be due to the primarily higher incidence of infection in developing countries and to the fact that half of the study participants stemmed from rural areas like in the current study area. Moreover, these patients are more prone to infection at the time of active disease owing to loss of immunoglobulin G, systemic factor B in urine, and T-lymphocyte cell dysfunction. This might be due to the activation of podocyte express receptors by the respective cytokines (IL-4 and IL-13) and may act on monocytes to produce vascular permeability factors involved in the pathogenesis of proteinuria in patients with relapse (10, 31).

Compared to children with normal nutritional status, wasting children were observed to have 1.93 times increased hazard of developing a relapse. This finding was supported by the study done in Ethiopia (14) and Indonesia (32). The possible explanation for this similarity might be due to social factors such as delayed early access to dietary intervention, lower socioeconomic status, and ignorance, which may affect the nutritional status of the patient with nephrotic syndrome. This could be explained by the podocyte biology theory of proteinuria and T-lymphocyte cell-mediated impairment, which lead to a relapse of nephrotic syndrome (33). According to the current study findings, approximately 93% of children who developed relapse had wasted and low serum albumin and protein levels, which supports the above justification.

This study also showed that a low serum albumin level was found to be a significant predictor of relapse among children with nephrotic syndrome. Accordingly, children who had an albumin level less ≤ 1.5 g/dl increased the hazard of relapse by 4.34 times as compared to those who had serum albumin levels ≥ 1.5 g/dl by adjusting the effect of other variables. This is in accordance with previous studies done in Ethiopia (14, 15), and Dhaka Medical College Hospital (DMCH) in Bangladesh (17). The possible explanation for the similarity might be due to the predominance of malnutrition in developing countries, which could create a favorable environment for the loss of albumin through urine. Because albumin is the main protein in the blood, it regulates oncotic pressure, preventing fluid from leaking into the extracellular space and causing edema (34). In line with this concept, more than half of children who had a serum albumin level less than or equal to 1.5 g/dl in the present study developed relapse.

Triglyceride level during previous remission was an independent predictor of relapse in nephrotic patients. As other variables remained constant, children with high serum triglyceride levels at remission had a 2.74 times greater hazard of relapse than those with a serum triglyceride level below 110 mg/dl. It was in agreement with the studies done in Bangladesh (44%) (35), India (56%) (36), and Ethiopia (88%), (14), which showed that elevated triglyceride levels during remission were a predictor of relapse. Congruently, in this study, 52% of children with elevated triglyceride levels experienced relapse. This could be explained by the podocyte biology theory of proteinuria pathogenesis that the elevated triglyceride level may alter the glomerular filtration barrier. Furthermore, an increased synthesis or decreased clearance of lipoprotein may contribute to elevated triglyceride levels and thus lead to a relapse of nephrotic syndrome (33).

## Limitations of the study

The findings of this study might suffer from the fact that it is a retrospective study and based on secondary data; some variables like socioeconomic status, parental educational status, and essential biochemical-related variables were missing, while the others were not recordable. Therefore, the study failed to track the relapse that occurred in those who had incomplete records and this may underestimate the relapse rate because patients excluded with incomplete medical records might be at high risk of complications and relapse. Moreover, since the majority of the observations were censored, there may be a potential bias due to excluded records and the unknown status of absconders.

## Conclusion

Relapse is a significant public health problem in children with nephrotic syndrome. The incidence of relapse among children with nephrotic syndrome remains high during the follow-up period. Children with wasting malnutrition, acute respiratory tract infections, elevated triglyceride levels, and low serum albumin levels are more likely to relapse.

## Recommendations

Healthcare providers should give special emphasis to nephrotic syndrome patients with acute respiratory tract infection and malnutrition at admission and follow patients’ biochemical-related findings attentively. The zonal health bureau and other stakeholders should improve the capacity of healthcare providers. A prospective study design would better compensate for the limitations of this study.

## Data Availability

The original data for this study are available from the corresponding author upon reasonable request.

## Acknowledgments

We would like to thank data collectors, supervisor, hospital staffs, and administrators for their unreserved efforts and commitment. Lastly. Our gratitude goes to Assosa University for funding this study and University of Gondar to pursue and facilitating this study.

